# Excess mortality during the COVID-19 pandemic in Aden governorate, Yemen: a geospatial and statistical analysis

**DOI:** 10.1101/2020.10.27.20216366

**Authors:** Emilie Koum Besson, Andy Norris, Abdulla S. Bin Ghouth, Terri Freemantle, Mervat Alhaffar, Yolanda Vazquez, Chris Reeve, Patrick J. Curran, Francesco Checchi

## Abstract

**Background:** The burden of COVID-19 in low-income and conflict-affected countries is still unclear, largely reflecting low testing rates. In parts of Yemen, reports indicated a peak in hospital admissions and burials during May-June 2020. To estimate excess mortality during the epidemic period, we quantified activity across all identifiable cemeteries within Aden governorate in Yemen (population approximately one million) by analysing very high-resolution satellite imagery, and compared estimates to Civil Registry office records from the city.

**Methods:** After identifying active cemeteries through remote and ground information, we applied geospatial analysis techniques to manually identify new grave plots and measure changes in burial surface area over a period from July 2016 to September 2020. After imputing missing grave counts using surface area data, we used alternative approaches, including simple interpolation and a generalised additive mixed growth model, to predict both actual and counterfactual (no epidemic) burial rates by cemetery and across the governorate during the most likely period of COVID-19 excess mortality (from 1 April 2020), and thereby compute excess burials. We also analysed death notifications to the Civil Registry office during April-July 2020 and in previous years.

**Results:** We collected 78 observations from 11 cemeteries, of which 10 required imputation from burial surface area. Cemeteries ranged in starting size from 0 to 6866 graves. In all but one a peak in daily burial rates was evident from April to July 2020. Interpolation and mixed model methods estimated ≈ 1500 excess burials up to 6 July, and 2120 up to 19 September, corresponding to a peak weekly increase of 230% from the counterfactual. Satellite imagery estimates were generally lower than Civil Registry data, which indicated a peak 1823 deaths in May alone. However, both sources suggested the epidemic had waned by September 2020.

**Discussion:** To our knowledge this is the first instance of satellite imagery being used for population mortality estimation. Findings suggest a substantial, under-ascertained impact of COVID-19 in this urban Yemeni governorate, and are broadly in line with previous mathematical modelling predictions, though our method cannot distinguish direct from indirect virus deaths. Satellite imagery burial analysis appears a promising novel approach for monitoring epidemics and other crisis impacts, particularly where ground data are difficult to collect.

## Introduction

On 10 April 2020, Yemen recorded its first laboratory-confirmed coronavirus disease (COVID-19) case in the Southern governorate of Hadhramout. By the end of May, cases and deaths had been reported in the governorates of Aden, Taiz, Lahj and Sanaa.^1^ The pandemic’s impact on Yemen was expected to be severe, despite its comparatively young population.^2,3^ Transmission models parameterised based on China and Europe evidence initially predicted some 85,000 virus deaths, approaching the number reported killed during the country’s ongoing armed conflict.^4–6^ Additional risk factors of concern resulting from this five-year protracted crisis included disrupted health services (only half of pre-existing health facilities were fully functional as of early 2020), shortage of health workers, overcrowding due to internal displacement, food insecurity and shrinking humanitarian aid.^3,5,7–9^

During May 2020, videos posted on social media showing large numbers of fresh graves suggested a spike in burial activity across several Yemeni governorates including Sana’a, Aden, Ibb and Al Baydha.^10–12^ In the southern city of Aden, the use of mechanical excavators in place of human gravediggers suggested that demand exceeded routine capacity.^13,14^ The medical organisation Médecins sans Frontières also reported a peak in hospital admissions, with a high case-fatality ratio, in Aden during the same period.^10^ Separately, a shortage of personal protective equipment forced several hospitals to close or reject patients exhibiting COVID-19 symptoms.^3,12,15^

As of 9 October 2020, Yemen had reported 2049 confirmed SARS-CoV-2 infections with 593 deaths^16^, but the reliability of these official numbers is undermined by low testing capability.^17,18^ A high COVID-19 case-fatality ratio (28.9%) is indicative of case under-ascertainment.^19,20^ The virus could thus be circulating undetected and unmitigated within communities, and the true mortality figures could be far higher than reported, as suggested elsewhere.^21–23^ Moreover, in Yemen’s protracted crisis an epidemic could result in increased population mortality both directly (from SARS-CoV-2 infection) and indirectly (due to reduced access to healthcare, lack of healthcare workers and/or medical supplies, and/or the socio-economic costs of the pandemic and related control measures, including worsened food security). Information on excess deaths can inform the ongoing response and provide evidence for additional resource mobilisation.^24^ Moreover, it may be combined with transmission-dynamic modelling to infer the local state of progression of the epidemic.^21^

In this study, we used a novel method based on analysis of satellite imagery to remotely estimate excess mortality during a COVID-19 epidemic in Aden, Yemen. Our study had the secondary aim of establishing the method for possible application elsewhere.

## Methods

### Study population and period

Our analysis covered the entire Aden governorate (population ≈ 1 million: see below), consisting mainly of urban or peri-urban settlements. We systematically measured burial activity over (i) a baseline pre-pandemic period starting in Jan 2016 and (ii) the COVID-19 transmission period, starting 1 Apr 2020 (see below) until mid-September 2020.

### Study design

We sought to exhaustively quantify the number of graves over time across the governorate, and compare burial rates during the two analysis periods. After gathering qualitative information on local burial practices, we identified all recognised burial sites with any activity since Jan 2016 and quantified new graves over time increments bounded by the availability of successive very high-resolution (VHR) satellite images, either through visual annotation of individual features (burial plots) or inference from graveyard surface area changes. We used longitudinal statistical analysis techniques to model burial rates in the baseline and pandemic period, either (i) for each burial site alone or (ii) using the entire dataset. We then projected the baseline rate to the pandemic period as a counterfactual, and used this to estimate excess deaths attributable to COVID-19. We also estimated population death rates using available demographic estimates. For comparison, we also present data on deaths notified to the Aden Civil Registry office, collected as part of a separate study.

### Data sources

#### List of cemeteries

We first gathered informal information on burial practices in Yemen, including during the COVID-19 epidemic, from a network of researchers, civil society actors and the British Yemeni diaspora, that we interacted with as part of separate projects. We also leveraged this network to ground-truth imagery analysis (see below).

To identify all burial sites within Aden governorate that may have been active during the study period, we relied on the above network, geospatial datasets and gazetteers such as OpenStreetMap^25^ and Google Maps^26^ and visual inspection of VHR imagery to create a master list of burial sites. We mapped the boundaries of burial sites and noted salient characteristics including alternative names, burial patterns and specific communities said to use them. Across the governorate, 27 burial sites were identified. After triangulating all available data sources, we were able to classify with reasonable certainty 11 sites as active and 16 as closed or featuring no/very few new burials across the study period.

#### Satellite imagery

We sourced the most suitable available VHR images for each cemetery during the analysis period, ensuring they were cloud-free, of high radiometric quality and most importantly, focussing on those with a spatial resolution between ∼31cm to ∼40cm per pixel. The higher resolution of 31cm was preferred where available, as it provided the best clarity of features on the ground and ensured the most accurate identification of new burials. We used coarser resolution imagery (∼40cm to ∼50cm) where no higher resolution alternative was available, to investigate its uses and limitations and, at a minimum, measure surface area changes (below).

Images were obtained as Ortho Natural Colour via SecureWatch (https://www.maxar.com/products/securewatch) and were delivered pre-processed, corrected for illumination and geometric distortion, and pan-sharpened. To further improve the detection of individual burial plots, some were enhanced using Edge Detection techniques (Sobel and/or Touzi filter, using the Orfeo ToolBox^27^).

#### Civil Registry records

Available records of burial permit requests to the Civil Registry offices of Aden governorate were tallied, covering the months of January to July 2020, and, for comparison, January to May in 2017, 2018 and 2019. Weekly records were available for weeks starting 4 May to 1 August 2020.

#### Armed conflict intensity data

We sourced armed conflict data compiled by the Armed Conflict Location & Event Data Project (ACLED), which has carried out intensive collection and curation of media and local civil society reports of insecurity events and fatalities since 2015, in partnership with the Yemen Data Project.^28^ The dataset contained 2315 individual event records; we computed daily event and fatality rates for the sub-district (administrative level 3) within which each cemetery is located, and for all other sub-districts combined.

#### Population denominators

We computed population estimates for Aden governorate so as to calculate per-population death rates. Alternative base sources included (i) Yemen Central Statistical Organisation (CSO) projections of June 2004 census data^29^, assuming a 2.78% annual increase for 2015-2020^30^; (ii) WorldPop annual estimates, built on a predictive model that redistributes total census population into ∼100 m^2^ grids based on relative population density^31^; and (iii) Facebook Data for Good estimates^32^, which combine machine learning analysis of satellite imagery with local census and household survey data. We averaged the three after weighting them based on a quality score computed from a published checklist.^33^ In 2016-17 and 2019 respectively, the International Organisation for Migration and the United Nations World Food Programme published estimates based on CSO projections but adjusted for crisis-related forced displacement: we applied the ratio of displacement-adjusted and CSO estimates to our weighted average to come up with a high-end denominator, which we used for analysis. Lastly, we smoothed the series to obtain monthly values (Figure S7).

### Imagery analysis

We used geospatial analysis techniques to remotely count the number of new individual burials (e.g. headstones) within cemeteries when visible and/or delineate the change in total burial area over time. We relied on multispectral images and edge detection to manually identify and annotate cemetery features. Trained analysts tagged individual burials using QGIS open source software.^34^ A total of 128,213 features were tagged across identified sites. Where possible, we used publicly available photographs and videos from media reports to validate our grave counts, as exemplified in Figure 1. To ensure the accuracy of annotations, all features have been quality-checked by two other analysts.

**Figure 1.**
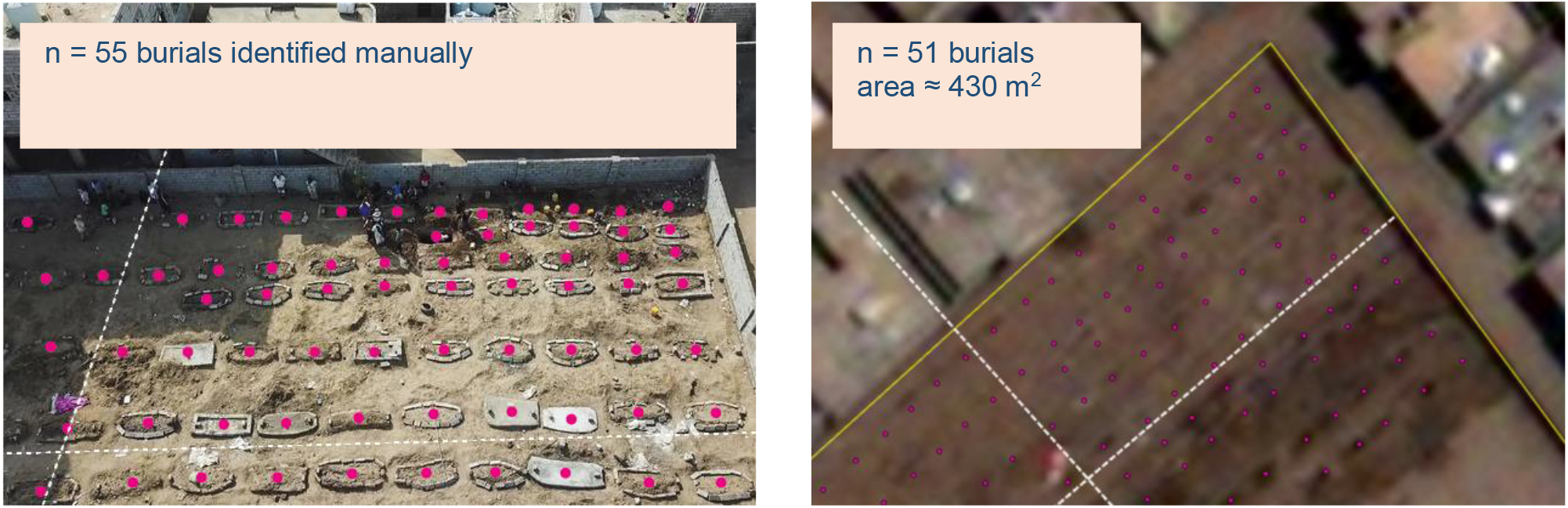
Publicly available media report drone photograph^35^ © Associated Press (left-hand panel) of a section of Radwan cemetery, Aden, and corresponding VHR satellite image view © Maxar Technologies 2020 (right-hand panel), both from 21 May 2020.

### Statistical analysis

While most of the available images featured sufficient resolution (30-35 cm per pixel) to identify individual graves, some (10/78, 12.8%) only had sufficient resolution (40-60 cm) to quantify expansions in surface area since the previous image time point. We thus imputed the missing grave counts through a predictive model trained on complete (both grave count and surface area) observations. Specifically, we fitted a quasi-Poisson generalised linear mixed model to the number of new graves since the previous image, with the natural log of new surface area as predictor, baseline grave count at the start of the time series as an offset, and cemetery as random effect. To avoid data sparsity, we did not partition data into training and holdout (validation) sets, but rather used leave-one-out cross-validation (LOOCV) and the Akaike Information Criterion^36^ to quantify out-of-sample predictive power.

Given the timing of marked peaks in daily burial rates observed across most cemeteries (Figure 3), we settled on 1 April 2020 as the most plausible start of the COVID-19 epidemic period in Aden governorate (this date is merely functional to our analysis, as detailed below, and is broadly interpretable as the earliest point at which COVID-19 attributable mortality would have caused a statistically observable excess: it is not to be mistaken with the date of virus introduction into Yemen). To estimate total and excess mortality during this period, we implemented two alternative approaches, as follows.

**Figure 2.**
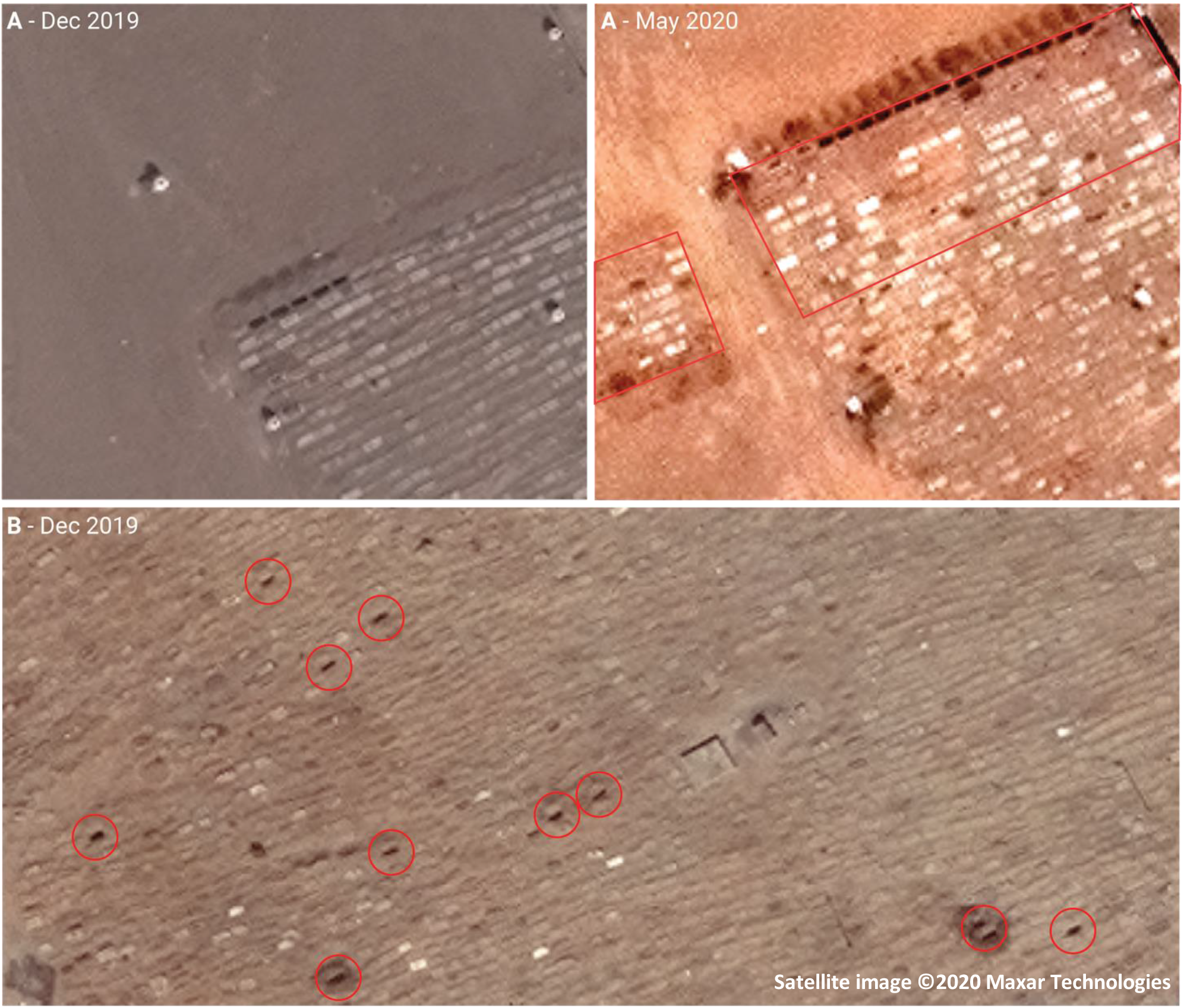
Sample of very high-resolution images from two cemeteries in Aden governorate, exemplifying the two typologies or burial pattern observed: (A) expansion into new ‘blocks’ (denoted by red outline) and (B) ‘infilling’ within existing burial area (denoted by red circles) Satellite imagery © Maxar Technologies.

**Figure 3.**
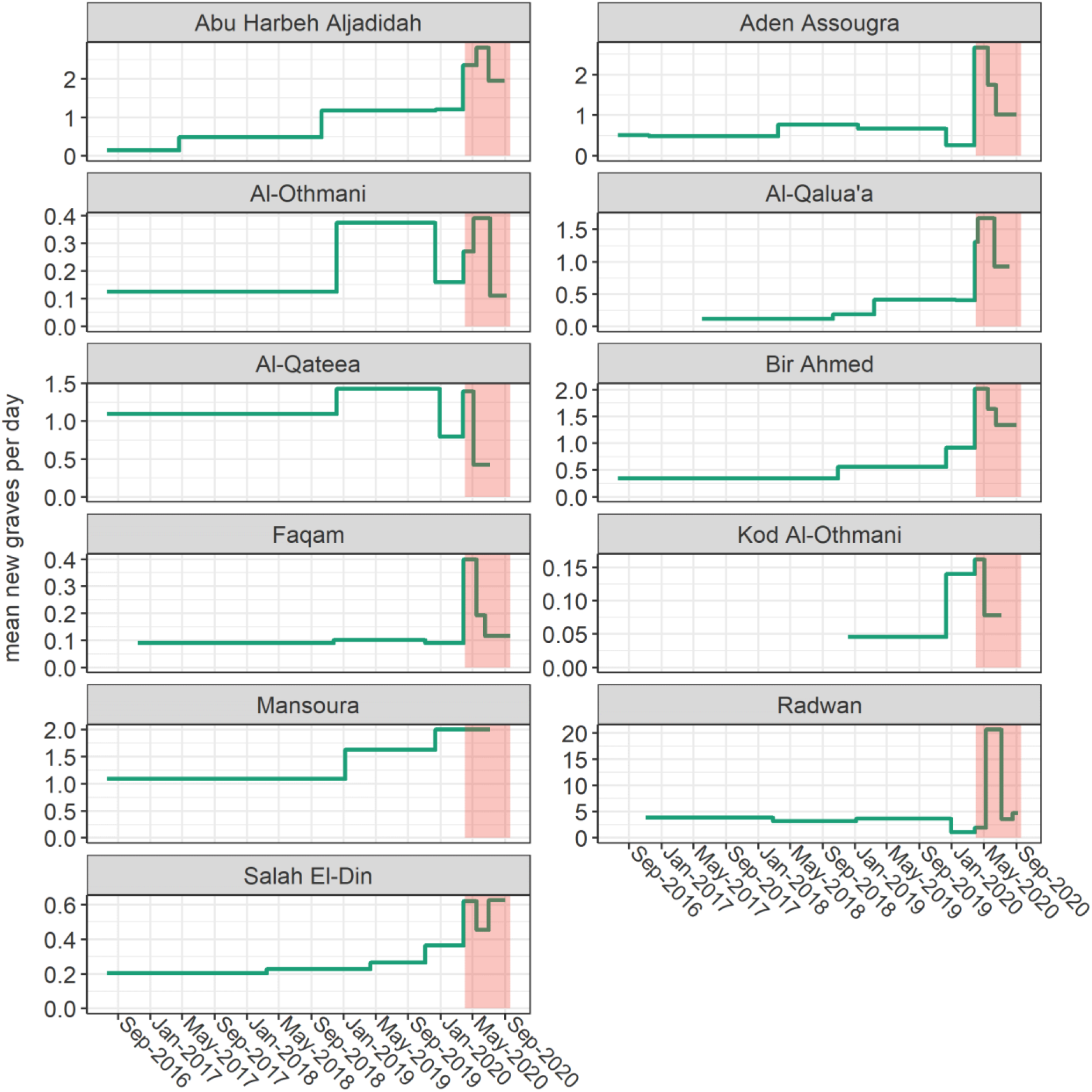
Evolution of burial rate over the analysis period, by cemetery. Each horizontal segment comprises the timespan between consecutive images. The epidemic period (1 April 2020 onwards) is shaded in red. Note that the y-axis scales are cemetery-specific.

We firstly did a ‘case-based’ analysis of each cemetery: this consisted of interpolating cumulative grave counts between each observation through smooth splines or, for comparison, simple linear segments.^37^ As this approach rests solely on information from the cemetery itself, we refrained from any extrapolation beyond the timespan of data availability; furthermore, to provide governorate-wide estimates we censored analysis at the latest time point for which an interpolated datum was available across all 11 cemeteries (6 Jul 2020: see Table 1). To calculate a counterfactual (i.e. burials in the absence of COVID-19) for each cemetery, we forward-projected the mean daily burial rate during the pre-epidemic (baseline) period (i.e. prior to April 2020) into the epidemic period, with a small upward correction reflecting population increase since the baseline. We then subtracted this counterfactual cumulative burial number from the actual estimated number to yield the excess and related statistics (e.g. burial rates per 1000 person-years), which we lastly summed across all cemeteries. We do not believe that this approach supports meaningful confidence interval calculation, and as such present only point estimates.

**Table 1.** Availability of imagery and characteristics of cemeteries included in the analysis.

| Cemetery | Sub-district | Number of observations | Timespan of available imagery |  | Number of graves |  | Surface area (m<sup>2</sup>) |  |
| --- | --- | --- | --- | --- | --- | --- | --- | --- |
|  |  |  | earliest | latest | starting | ending | starting | ending |
| Abu Harbeh Aljadidah | Al Mansurah | 8 | 21/07/2016 | 29/08/2020 | 0 | 1289 | 0 | 6088 |
| Aden Al-soughra | Al Burayqah | 9 | 21/07/2016 | 01/09/2020 | 222 | 1265 | 2797 | 11,821 |
| Al-Othmani | Ash Shaykh Othman | 7 | 21/07/2016 | 07/09/2020 | 261 | 568 | n/a | n/a |
| Al-Qalua'a | Dar Sad | 8 | 02/06/2017 | 06/08/2020 | 0 | 415 | 0 | 2368 |
|  |  |  | earliest | latest | starting | ending | starting | ending |
| Al-Qateea' | Kritar - Sirah | 6 | 21/07/2016 | 06/07/2020 | 6866 | 8522 | n/a | n/a |
| Bir Ahmed | Al Burayqah | 7 | 21/07/2016 | 01/09/2020 | 1039 | 1909 | 24,292 | 31,735 |
| Faqam | Al Burayqah | 7 | 15/11/2016 | 19/09/2020 | 777 | 929 | n/a | n/a |
| Kod Al-Othmani | Ash Shaykh Othman | 5 | 05/12/2018 | 06/07/2020 | 212 | 255 | n/a | n/a |
| Al-Mansoura | Al Mansurah | 4 | 21/07/2016 | 06/07/2020 | 2010 | 3960 | n/a† | n/a |
| Al=Radwan | Ash Shaykh Othman | 9 | 03/11/2016 | 07/09/2020 | 660 | 6507 | 4035 | 38,954 |
| Salah Al-Deen | Al Burayqah | 8 | 21/07/2016 | 01/09/2020 | 755 | 1167 | 5580 | 9198 |

† n/a indicates that the cemetery has not expanded in orderly 'blocks' of new area, but rather that new graves were added to existing burial areas in an 'infilling' pattern: in such cases, we did not consider surface area a meaningful metric.

Secondly, we fitted a predictive model to data from all cemeteries, and used this to infer burials across the timespan of imagery availability for any cemetery (21 July 2016 to 19 September 2020). As the observed pattern of burials was non-linear, and after exploring various linear options (least-squares, generalised linear or quadratic count models), we settled on a generalised additive model for location, scale and shape (GAMLSS)^38^, fitted using the R `gamlss` package; this framework allows for fitting non-linear smoothing functions and estimating both the mean and the shape of distributions of the dependent variable. We assumed a quasi-Poisson distribution for the cumulative number of graves since the start of each cemetery time series, with time (day) since the start of the analysis period as a ‘baseline’ growth predictor and time since the start of the epidemic period as a further ‘added growth’ predictor;^39^ time was nested within cemetery as a random effect term to capture unmeasured factors explaining cemetery-specific growth trajectories. We used penalised B-splines of time^40^ as additive smoothing terms to estimate both the mean and the shape (overdispersion) of the distribution of graves. We selected the model through AIC and visual inspection of the observed fit, including model assumption diagnostics. We then used the model to predict the actual number of graves across all cemeteries, as well as a counterfactual (specified by setting epidemic time to zero), with the difference representing the excess. As GAMLSS models do not yield accurate standard errors, we computed prediction confidence intervals by posterior simulation from the model coefficient values. Below we also present an alternative model including the rate of insecurity events within the cemetery’s sub-district as a further fixed effect, so as to adjust for conflict-related mortality changes; the addition of this predictor did not significantly improve model fit. Statistical analysis was done in R^41^. Data and analysis code are accessible at https://github.com/francescochecchi/aden_covid, and we welcome alternative analysis suggestions.

### Ethics

The satellite imagery study was approved by the Ethics Committee of the London School of Hygiene & Tropical Medicine (ref. 22458). The Ministry of Health in Aden was notified of the former and approved collection of Civil Registry data. No identifiable personal data were used, and satellite images were not of sufficient resolution to identify individual people.

## Results

### Description of burial patterns

Table 1 summarises data availability and characteristics of the 11 cemeteries across Aden governorate for which available imagery indicated any activity between 2016 and September 2020. Six were within the city itself; five appeared to add graves solely by allocating new ‘blocks’, while the remainder partly or only adopted an ‘infilling’ pattern: these distinct typologies are illustrated in Figure 2. On average, 7.1 images were available per cemetery, for a total 78 observations. Imputation of grave counts from surface area was reasonably accurate (Figure S9), and the model did not lose predictive power on cross-validation (Table S4).

As shown in Figure 3, the trajectory of daily burial rates differed considerably across cemeteries; however, in nearly all, a peak during April-July 2020 was visible, with evidence of decline in some during August-September.

### Excess burial estimates

Summary estimates from the case-based analysis approach are shown in Table 2; results by cemetery are in Table S5 and Table S6. Smooth spline and linear interpolation yielded very similar estimates for the period 1 April to 6 July 2020 (96 days). Around 1500 excess deaths are estimated to have occurred during this period, corresponding to standardised mortality ratios of 2.68 (smooth spline) and 2.81 when comparing the actual burial rate to the projected counterfactual.

**Table 2.** Estimates of baseline, epidemic-period and excess burials across Aden governorate, by analysis approach.

| Statistic | Case-based analysis<br>1 Apr to 6 Jul 2020 |  | GAMLSS model<br>Estimate (95%CI) |  |
| --- | --- | --- | --- | --- |
|  | Smooth spline | Linear | 1 Apr to 6 Jul 2020 | 1 Apr to 19 Sep 2020 |
| Baseline (counterfactual) burial rate<br>(per 1000 person-years) | 3.61 | 3.58 | 2.73 (2.57 to 2.87) | 2.78 (2.60 to 2.96) |
| Excess burial rate<br>(per 1000 person-years) | 5.41 | 5.82 | 6.24 (5.02 to 7.78) | 4.65 (0.93 to 9.08) |
| Total burials | 2309 | 2420 | 2320 (1904 to 2735) | 3424 (1554 to 5600) |
| Excess burials | 1451 | 1560 | 1598 (1285 to 1990) | 2120 (424 to 4137) |

Table 3 shows fit statistics for the GAMLSS model of all cemeteries. Model diagnostics are shown in Figure S12, suggesting moderate departures from the assumed normality of residuals. Visual inspection indicated reasonable fit to observed data points, though the model considerably under- or over-predicted burials during 2020 in two large cemeteries (Al-Radwan and Al-Qateea’: Figure S10, Figure S11). The model predicted a considerable increase in burials during April-July, with a resumption of the pre-epidemic rate of burial increase thereafter (Figure 4). Overall, the GAMLSS approach yielded similar estimates to case-based analysis for comparable periods (Table 2). By 19 September 2020, it predicted some 2100 excess deaths, though with a wide confidence interval. An alternative model featuring the rate of insecurity events within the cemetery’s sub-district as an additional predictor yielded 2413 excess deaths (95%CI 1427 to 3507); related data are in Table S8 and Figure S14.

**Table 3.** GAMLSS model fit statistics. Coefficients are exponentiated to provide linear scale rate ratios.

| Term | Rate ratio | p-value |
| --- | --- | --- |
| Penalised B-spline smoothing terms for the mean (expectation) parameter): |  |  |
| Baseline growth (day) | 1.004 | < 0.001 |
| Added epidemic growth (day) | 0.996 | < 0.001 |
| Penalised B-spline smoothing terms for the shape (overdispersion) parameter): |  |  |
| Baseline growth (day) | 0.991 | < 0.001 |
| Added epidemic growth (day) | 1.038 | < 0.001 |
|  | Akaike Information Criterion = 748.3 | Residual degrees of freedom = 48.1 |

**Figure 4.**
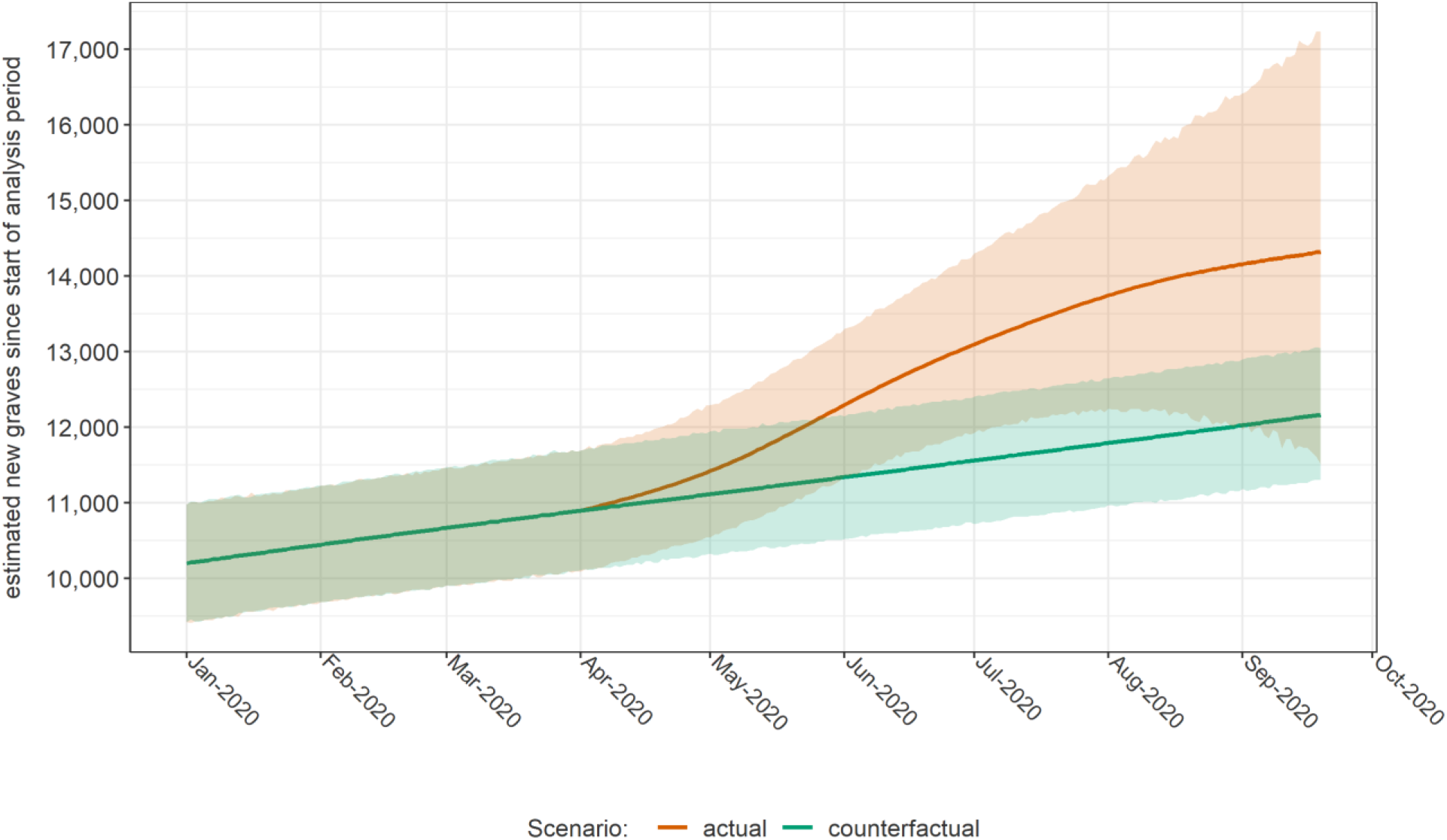
Evolution of the cumulative number of new graves across Aden governorate since the start of the analysis period, as estimated by the GAMLSS model, in actuality and in a counterfactual (no epidemic) scenario. Shaded areas indicate 95% confidence intervals. Only the year 2020 is shown.

Figure 5 and Figure S13 show, respectively, weekly (for 2020 only) and monthly comparisons of our statistical estimates with available records from the Civil Registry office: while patterns are consistent, the latter suggests a far steeper peak in death notifications in May 2020. During months before and after May, satellite imagery analysis yielded higher burial estimates; by contrast, Civil Registry records were consistently higher in earlier years, particularly in 2017 when satellite imagery estimated about half the number of burials as Civil Registry records (Figure S13). During a period (April to July 2020 inclusive) when the two sources were both available, GAMLSS and Civil Registry yielded 2834 and 3417 total deaths / burials (crude death rate 8.8 versus 10.6 per 1000 person-years), and 1940 and 2205 excess deaths (6.0 versus 6.8 per 1000 person-years), respectively (we used the median number of monthly death notifications in 2017-2019 as the counterfactual Civil Registry value).

**Figure 5.**
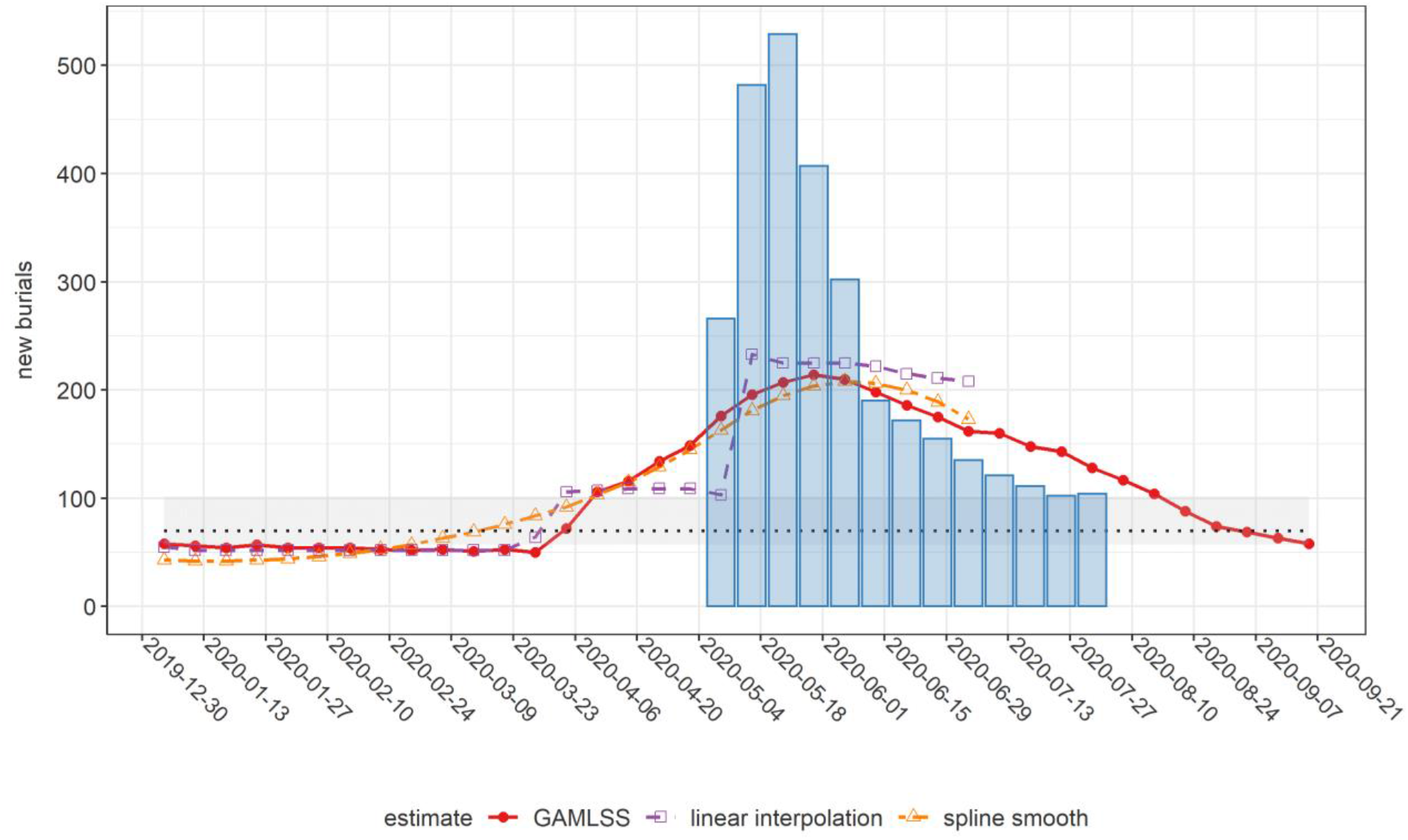
Comparison of weekly estimates of new burials across Aden governorate, by analysis method, and corresponding records from the Civil Registry office (blue columns), during 2020. The horizontal dotted line and shaded area indicate the median, minimum and maximum weekly burials based on the 2017-2019 Civil Registry time series.

## Discussion

This study offers evidence of considerable excess mortality temporally coinciding with COVID-19 transmission in Aden governorate, and suggests that at least the first wave of the epidemic in this urban part of Yemen had peaked and declined by September 2020. Both satellite imagery estimates and Civil Registry records point to considerable under-ascertainment of the true burden of COVID-19: only 34 COVID-19 deaths were officially reported in Aden over our analysis timespan.^16^

Mathematical modelling estimates had suggested that, without mitigation measures, Yemen could see some 58-84,000 deaths (or about 1.9 to 2.8 per 1000 persons) from COVID-19 disease during the first six months of the epidemic^4^. While this per capita prediction is broadly consistent with our estimate for Aden over a roughly similar timeframe (≈2.1, or ≈2.4 of the model with insecurity events is used), our analysis encompasses deaths both directly and indirectly (e.g. from other causes occurring in the context of overburdened health services and pandemic control measures: see below) attributable to COVID-19: during April 2020, severe floods affected Aden, and increased incidence of vector-mediated infections including malaria, dengue and chikungunya were noted in subsequent months. Moreover, it is plausible that SARS-CoV-2 has been more transmissible in Aden than in rural parts of Yemen due to greater social mixing, resulting in an earlier and more dramatic peak in the city: as such, our results should be extrapolated with caution to the rest of the country.

Given varyingly limited testing capacity and locally different approaches to defining and recording COVID-19 deaths, excess mortality may be the single most useful metric for establishing the full impact of the pandemic and drawing comparisons across geographical settings.^24,42^ This excess captures the “whole system” impact of the pandemic, as summarised in Figure 6. In Yemen, widely reported health facility closures to due lack of protective equipment, reduced confidence in health services and disruptions in humanitarian assistance could all have contributed substantially to increased non-COVID-19 mortality.

**Figure 6.**
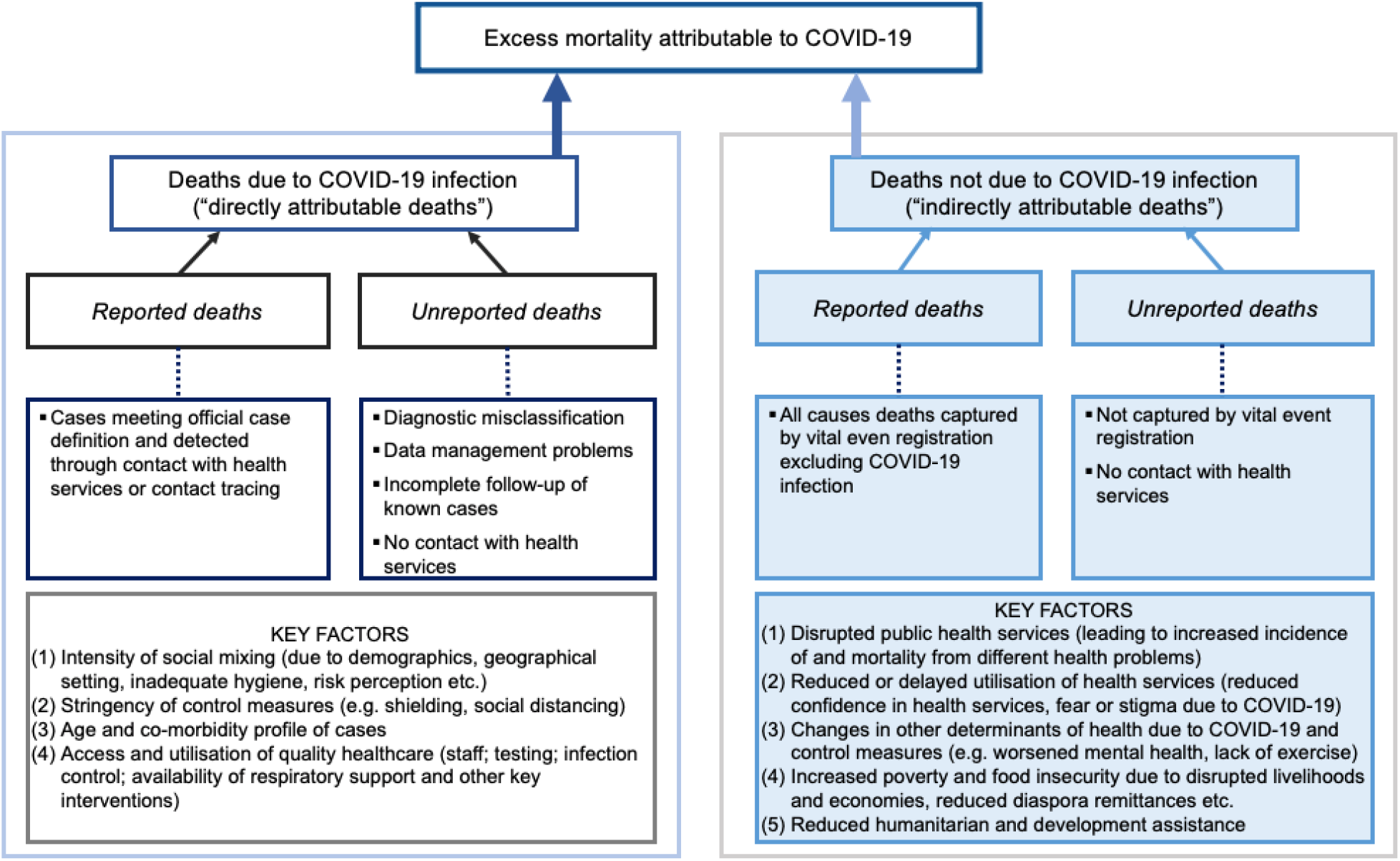
Diagram of direct and indirect contributions to excess COVID-19 mortality.

COVID-19 excess mortality evidence from low- and middle-income countries is only beginning to emerge. Cities in high-income countries experienced peak death rates some 200% (London) to 400% (Madrid) and 600% (New York) the baseline during March-April 2020.^43^ While our findings indicate similar-magnitude peaks in Aden (≈ 650% according to Civil Registry data, or ≈ 250% based on satellite imagery), at least three factors complicate this comparison: (i) Aden’s much younger population would, all else being equal, be expected to account for lower case-fatality ratios and thus per-capita mortality; (ii) Europe and North America mortality reflects scenarios of high access to respiratory support and intensive care, while in Aden this is likely to have been limited; and (iii) high-income countries mostly adopted strict and lengthy lockdown measures: by contrast, in Yemen and Aden, though school closures and restrictions on travel and public gatherings were reported^44^, measures appear to have been of shorter duration and, at least countrywide, population mobility seems no lower than pre-pandemic.^45^ Additional factors, such as the specific gender, age and co-morbidity profile of Aden’s population and travel during EidAl-Fitr celebrations (end May 2020), may have played a role.

Considerable under-ascertainment of COVID-19 deaths has been reported in Damascus^21^, Peru^46^ and various urban settings worldwide.^47,48^ While bureaucratic reasons (e.g. counting only laboratory-confirmed cases) and limited testing and health service access account for some of this gap, stigma, distrust of COVID-19 services, rumours surrounding the disease and other socio-cultural factors limiting service utilisation have been documented in Yemen.^49^ Taken together, these factors should fully be explored before concluding that the virus’ impact is adequately captured by formal surveillance statistics, or that its severity is milder in low-income countries.

### Study limitations

Problems with imagery availability and quality constrained the quantity of cemetery observations for analysis, reducing the accuracy of models and imposing limits on the period of analysis. Furthermore, the inherent inconsistencies (differences in viewing geometry, spatial and spectral resolution) of archive satellite imagery made it difficult to rapidly develop an automated, generalisable geospatial analysis method, and thus constrained us to a manual approach.

Comparison with Civil Registry data and, crudely, with the projected crude death rate in Yemen (around 6 per 1000 person-years^50^) suggests that imagery analysis under-estimated the true death toll, particularly in more remote periods (note however that Aden’s young, working population might in fact have a lower death rate than nationally). Plausible explanations could include (i) incomplete identification of active cemeteries (we could not access a formal list of cemeteries or a land registry against which to validate our list of 11 sites); (ii) problems identifying fresh graves in cemeteries where ‘infilling’ took place, largely due to insufficient VHR imagery; (iii) use of small, informal burial plots; and (iv) burial of decedents in ancestral areas of origin outside the governorate. The latter two phenomena are plausible when considering the reported spike in burial costs during the analysis period^13^, and the ongoing economic crisis affecting Yemenis. Uncertainty in population denominators could also bias estimated rates. By contrast, ground informants discounted other explanations such as mass graves and multiple bodies being buried together.

Lastly, our analysis is subject to causal inference limitations: we cannot distinguish direct from indirect mortality; the temporal association of SARS-CoV-2 transmission and excess mortality is not conclusive evidence that the pandemic caused the observed increase in burials; and, relatedly, it is inherently impossible to establish whether the counterfactual mortality trajectories we projected would indeed have borne out in reality.

### Conclusions and further work

Despite limitations, we believe this novel satellite imagery method holds promise and could become an efficient option for rapidly monitoring population mortality in settings without effective vital registration, particularly where ground access is arduous. Moreover, if the method were applied prospectively, and given sufficient funding, data availability and quality could be improved by commissioning bespoke imagery with the desired specifications, an option increasingly available given the deployment of new satellite sensors and increased control over imagery acquisition. Our manually analysed dataset also offers a platform for developing techniques including (but not limited to) feature extraction, change detection, segmentation, and supervised classification, and training a machine learning model to perform automated analysis. Automation would in turn increase the method’s efficiency and enable its rapid implementation in many locations at once. Increased use of geospatial analysis in crisis-affected settings has been advocated elsewhere.^51^

Our study helps to characterise the epidemic progression of COVID-19 in a low-income, crisis-affected context, and could help refine predictions of the virus’ future epidemiology in Yemen. Our excess mortality estimates allow international comparisons that can inform understanding of how different national strategies and policies have affected the spread and severity of the pandemic.

## Supporting information

Frequently Asked Questions

Abstract in Arabic

## Data Availability

The data and code files that support the findings of this study are openly available on githhub.com/francescochecchi.

https://github.com/francescochecchi/aden_covid

## Acknowledgements

We are grateful to colleagues at the United Kingdom Foreign, Commonwealth and Development Office, in particular Fergus McBean, for supporting this study. We also acknowledge the contribution of Anna Carnegie (LSHTM), Prof Huda Basaleem and Dr Khaled Zain (Faculty of Medicine and Health Sciences, University of Aden), the Yemen Analysis Hub at ACAPS, analysts and ground collaborators working with the Satellite Applications Catapult (including Benjamin Harris and Kazi Momin Ashraf), and organisations that shared population estimates and related information (the international Organization for Migration, the United Nations World Food Programme and Facebook Data for Good). Lastly, we are grateful to Dr Bothaina Attal, Dr Fouad Othman, Dr Ali Al-Mudhwahi and Dr Najwa Al-Dheeb for precious comments.

## Supplementary tables and figures

**Figure S7.**
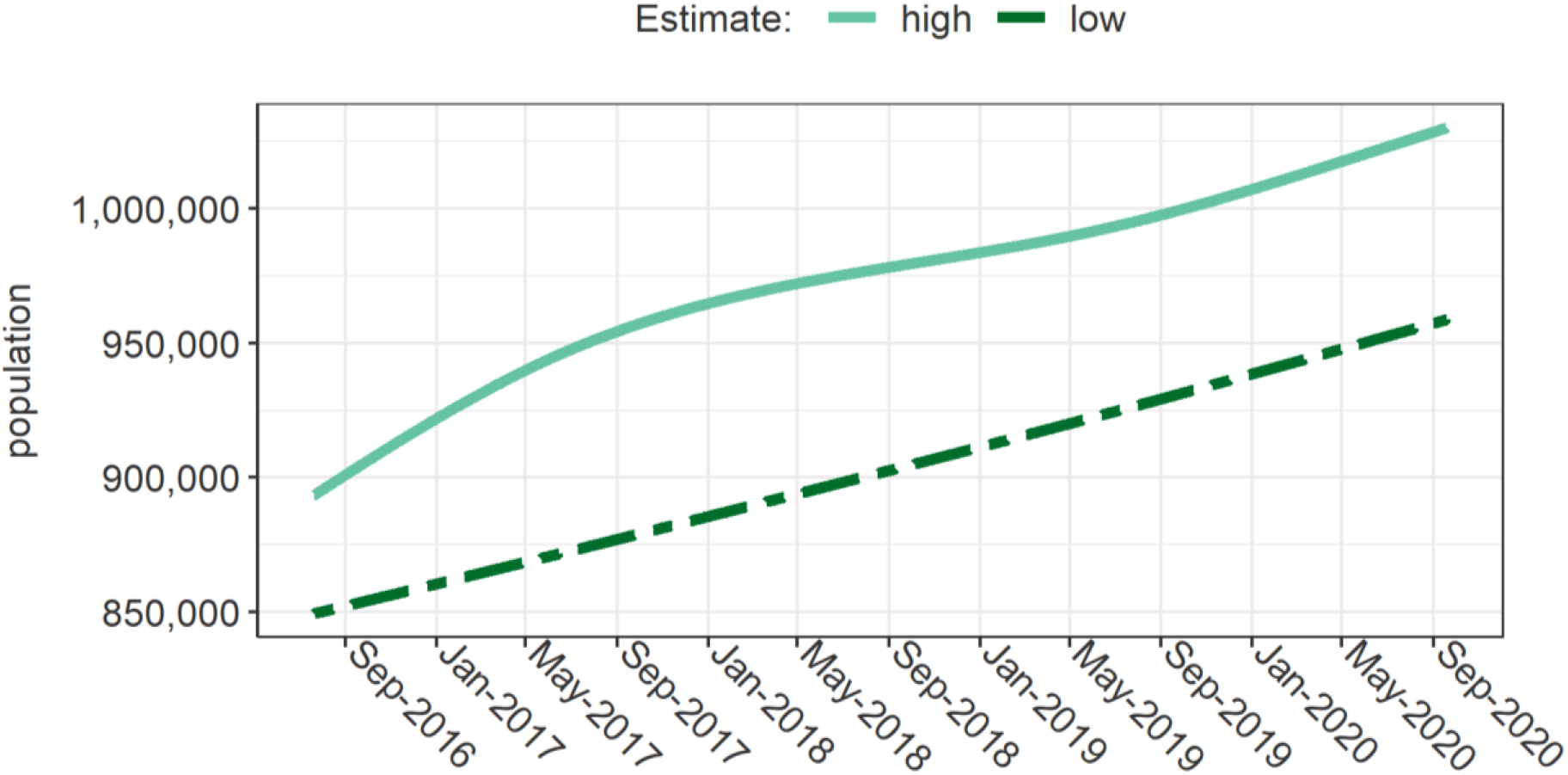
Estimated evolution of the population size of Aden governorate. The high-end estimate includes adjustment for displacement, and is adopted for the analysis. The low-end estimate is solely a weighted average of available base sources.

**Figure S8.**
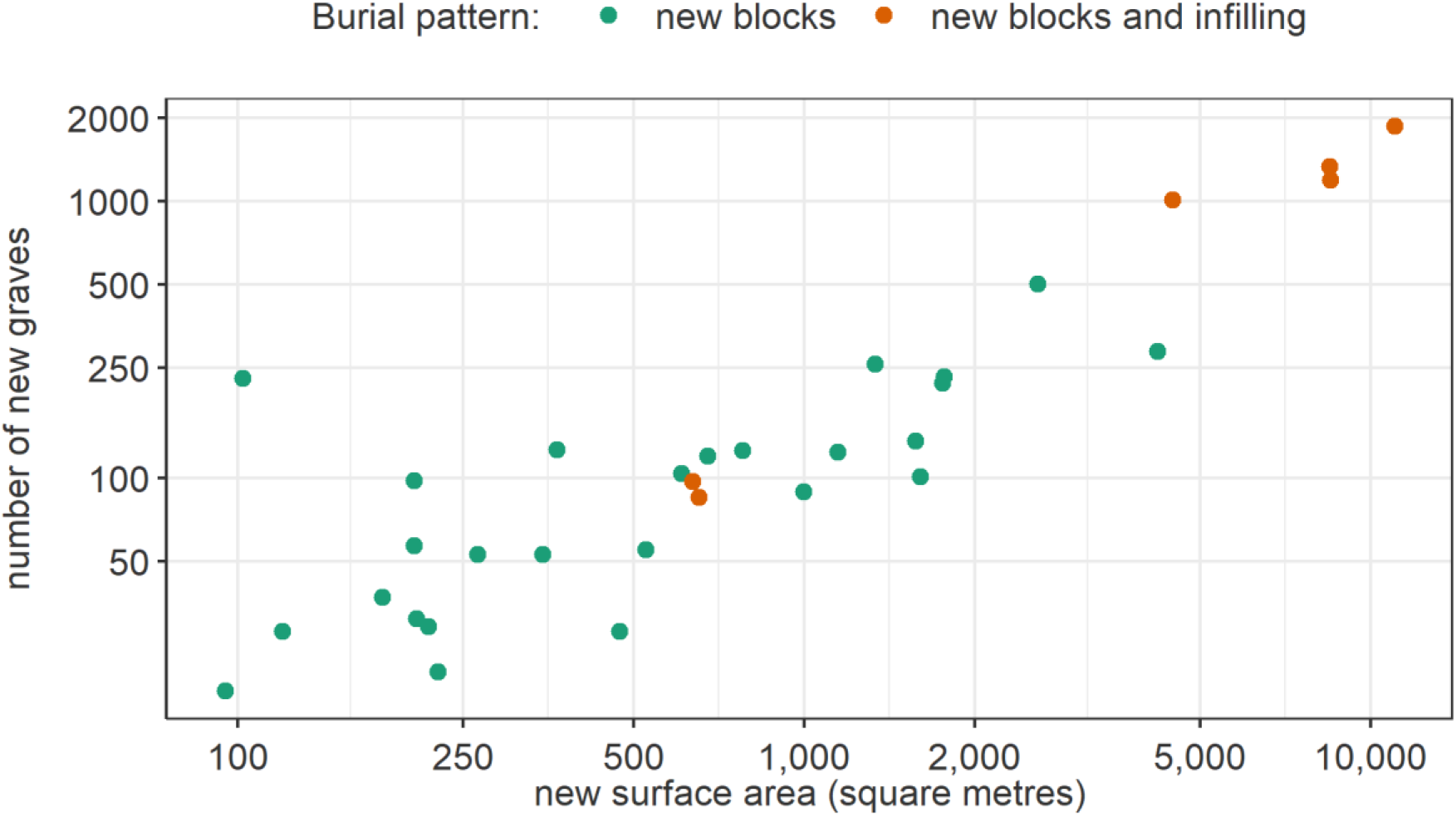
Correlation between the number of new graves and the new surface area expansion since the previous satellite image, by dominant burial pattern in the cemetery. Each plotted observation is an image (time point) within a given cemetery.

**Table S4.** Predictive model to impute new graves based on new surface area and baseline grave count. Only fixed effects are shown.

| Term | Coefficient | SE | p-value |
| --- | --- | --- | --- |
| intercept | -5.025 | 1.465 | < 0.001 |
| ln (new surface area in m<sup>2</sup>) | 0.821 | 0.079 | < 0.001 |
| AIC on training dataset: | 396.5 | Mean AIC across all LOOCV folds: | 385.1 |

SE: Standard error. AIC: Akaike Information Criterion. LOOCV: leave-one-out cross-validation.

**Figure S9.**
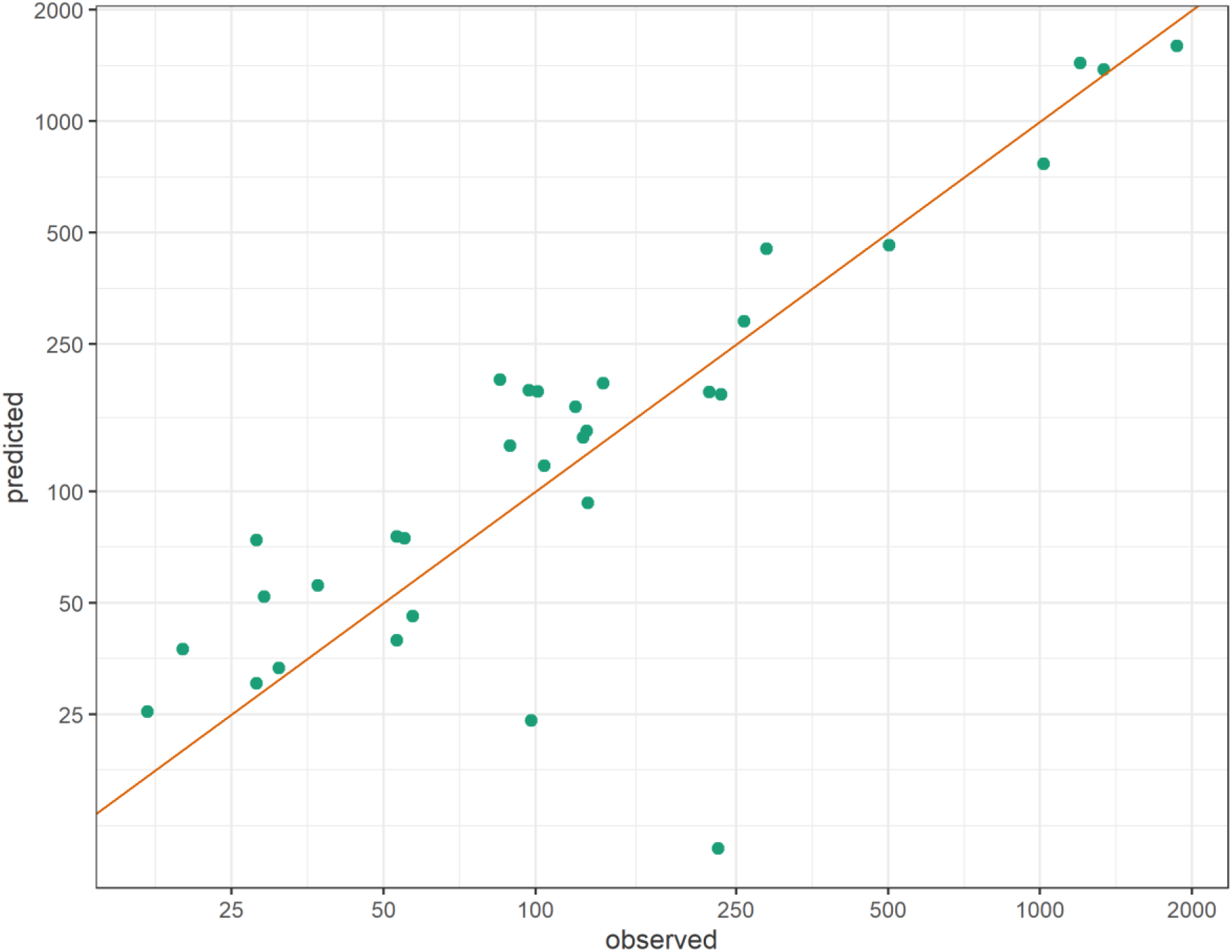
Predicted versus observed number of new graves after leave-one-out cross-validation. Each point indicates the observed and predicted values for the *k*^th^ fold (i.e. single observation) left out of the dataset, in turn. The red line indicates perfect agreement between observations and predictions.

**Table S5.** Estimates of baseline, epidemic period and excess burials by cemetery, based on smooth spline interpolation. Estimates of excess burials cover the period 1 April 2020 to 6 July 2020.

| Cemetery | Duration of periods (days) |  | Burial rate per day |  |  | Cumulative number of deaths |  |  |
| --- | --- | --- | --- | --- | --- | --- | --- | --- |
|  | baseline | epidemic | baseline | epidemic | excess | counterfactual | actual | excess |
| Bir Ahmed | 1350 | 96 | 0.46 | 1.76 | 1.27 | 47 | 169 | 122 |
| Al-Qalua'a | 1034 | 96 | 0.24 | 1.45 | 1.20 | 24 | 139 | 115 |
| Aden Al-sougra | 1350 | 96 | 0.59 | 1.98 | 1.36 | 60 | 190 | 130 |
| Al-Mansoura | 1350 | 96 | 1.30 | 1.99 | 0.62 | 132 | 191 | 59 |
| Abu Harbeh Aljadidah | 1350 | 96 | 0.70 | 2.48 | 1.74 | 70 | 238 | 167 |
| Kod Al-Othmani | 483 | 96 | 0.07 | 0.11 | 0.04 | 7 | 10 | 4 |
| Al-Othmani | 1350 | 96 | 0.20 | 0.33 | 0.12 | 20 | 31 | 11 |
| Al-Radwan | 1245 | 96 | 3.44 | 12.42 | 8.81 | 347 | 1192 | 846 |
| Salah Al-Deen | 1350 | 96 | 0.24 | 0.55 | 0.30 | 24 | 53 | 29 |
| Faqam | 1233 | 96 | 0.10 | 0.27 | 0.17 | 10 | 26 | 17 |
| Al-Qateea' | 1350 | 96 | 1.18 | 0.73 | -0.51 | 119 | 70 | -49 |

**Table S6.** Estimates of baseline, epidemic period and excess burials by cemetery, based on linear interpolation. Estimates of excess burials cover the period 1 April 2020 to 6 July 2020.

| Cemetery | Duration of periods (days) |  | Burial rate per day |  |  | Cumulative number of deaths |  |  |
| --- | --- | --- | --- | --- | --- | --- | --- | --- |
|  | baseline | epidemic | baseline | epidemic | excess | counterfactual | actual | excess |
| Bir Ahmed | 1350 | 96 | 0.46 | 1.78 | 1.29 | 47 | 171 | 124 |
| Al-Qalua'a | 1034 | 96 | 0.24 | 1.46 | 1.21 | 24 | 140 | 116 |
| Aden Al-sougra | 1350 | 96 | 0.58 | 2.05 | 1.44 | 59 | 197 | 138 |
| Al-Mansoura | 1350 | 96 | 1.30 | 2.02 | 0.64 | 132 | 194 | 62 |
| Abu Harbeh Aljadidah | 1350 | 96 | 0.69 | 2.56 | 1.83 | 70 | 246 | 176 |
| Kod Al-Othmani | 483 | 96 | 0.07 | 0.11 | 0.04 | 7 | 10 | 4 |
| Al-Othmani | 1350 | 96 | 0.20 | 0.35 | 0.15 | 20 | 34 | 14 |
| Al-Radwan | 1245 | 96 | 3.47 | 13.28 | 9.64 | 349 | 1275 | 926 |
| Salah Al-Deen | 1350 | 96 | 0.24 | 0.55 | 0.30 | 24 | 53 | 29 |
| Faqam | 1233 | 96 | 0.09 | 0.28 | 0.18 | 10 | 27 | 17 |
| Al-Qateea' | 1350 | 96 | 1.17 | 0.76 | -0.48 | 119 | 73 | -46 |

**Figure S10.**
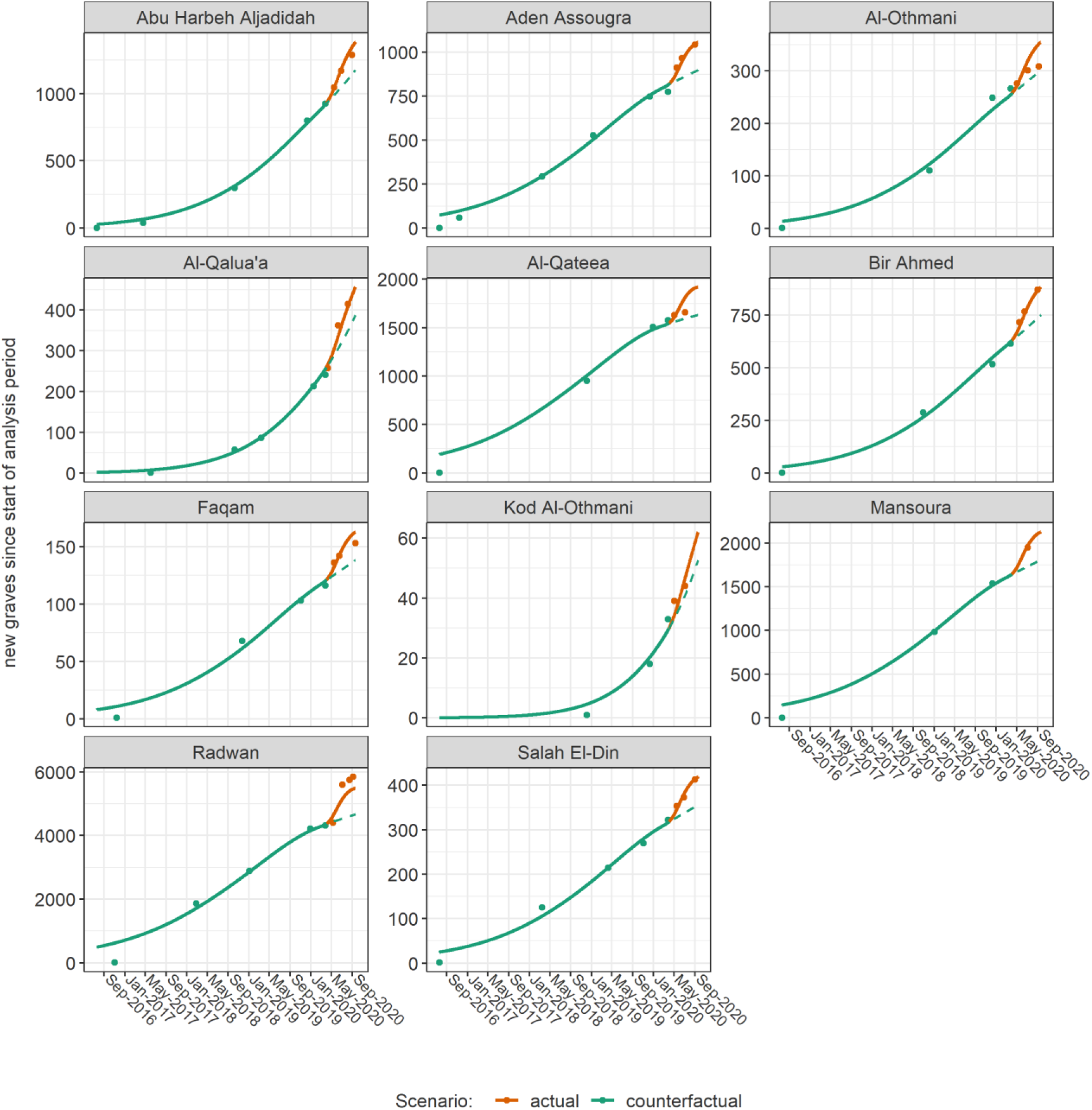
GAMLSS model predictions (lines) versus observations (dots), by cemetery. The dashed lines denote counterfactual predictions.

**Figure S11.**
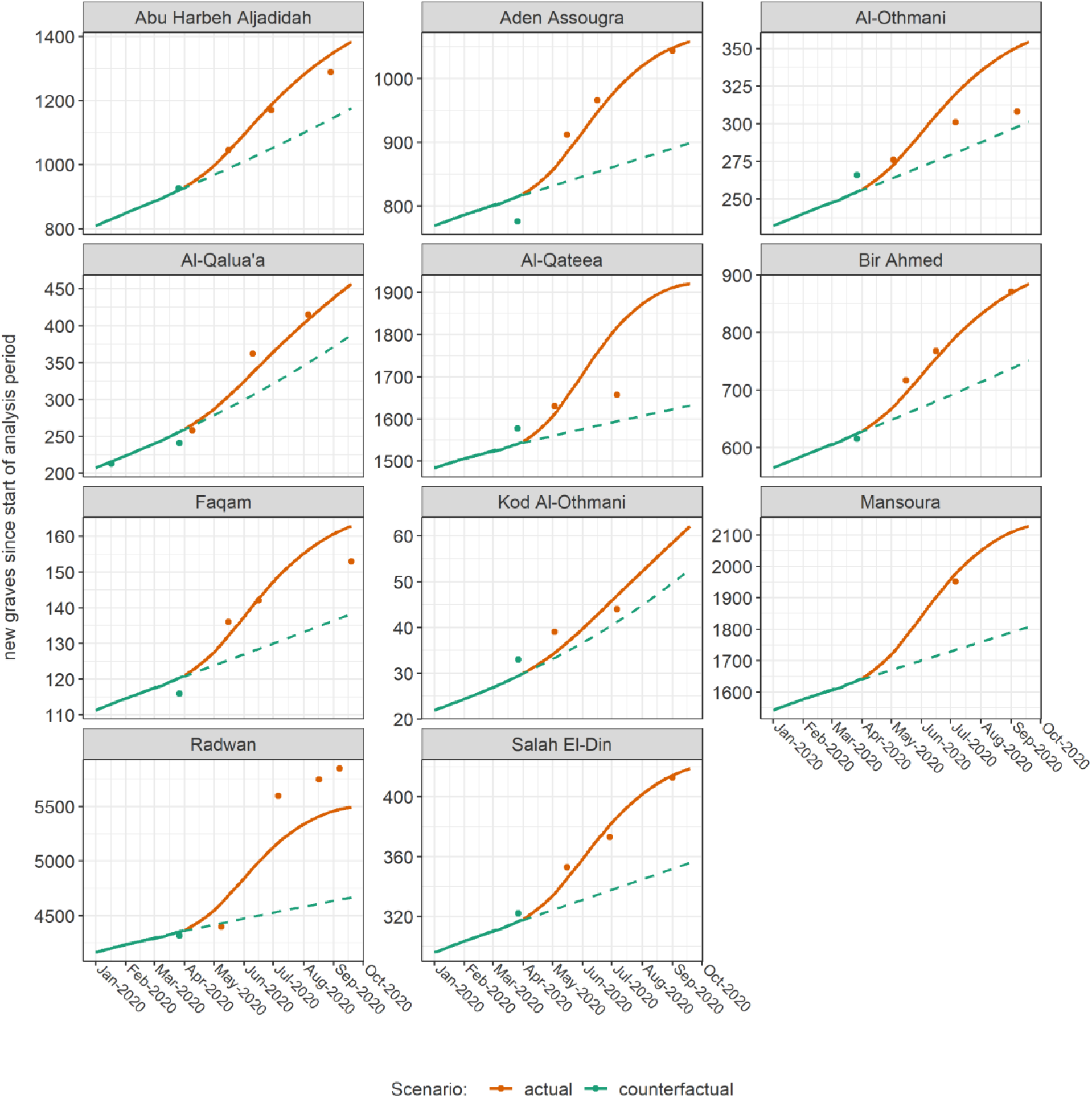
GAMLSS model predictions (lines) versus observations (dots), by cemetery, for the first 9 months of 2020. The dashed lines denote counterfactual predictions.

**Figure S12.**
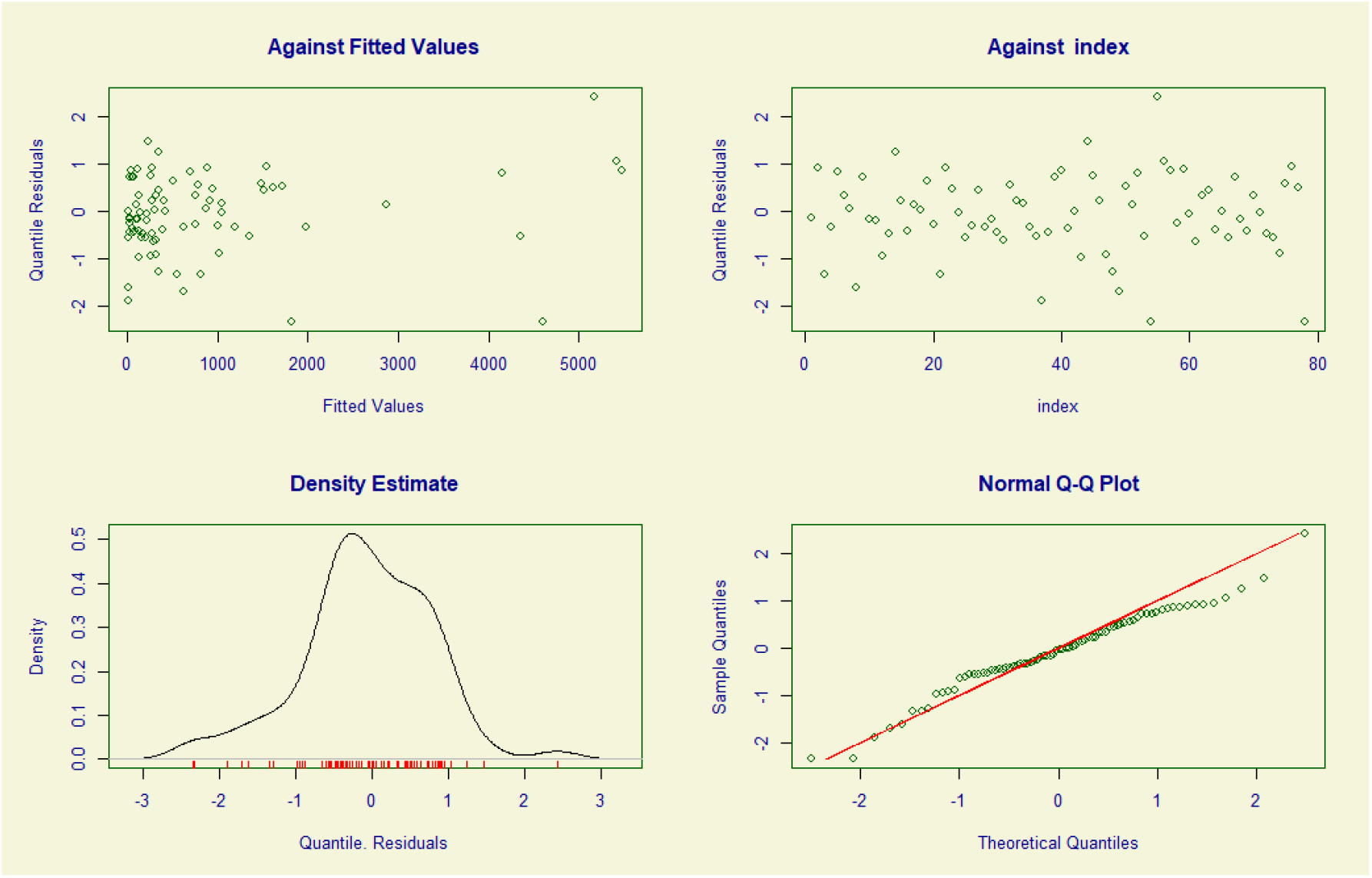
GAMLSS model diagnostic plots.

**Table S7.** Estimates of actual, counterfactual and excess burials by cemetery, based on the GAMLSS model. Estimates cover the period 1 April 2020 to 19 September 2020 (171 days).

| Cemetery | Burial rate per day (95%CI) |  |  | Cumulative number of deaths (95%CI) |  |  |
| --- | --- | --- | --- | --- | --- | --- |
|  | counterfactual | actual | excess | counterfactual | actual | excess |
| Bir Ahmed | 4.39<br>(4.04 to 4.80) | 5.17<br>(4.25 to 6.19) | 0.76<br>(0.10 to 1.54) | 751<br>(690 to 821) | 884<br>(726 to 1059) | 130<br>(17 to 263) |
| Al-Qalua'a | 2.27<br>(2.04 to 2.51) | 2.67<br>(2.12 to 3.23) | 0.41<br>(0.07 to 0.74) | 388<br>(349 to 429) | 456<br>(362 to 552) | 70<br>(12 to 127) |
| Aden Al-sougra | 5.26<br>(4.88 to 5.67) | 6.19<br>(4.99 to 7.49) | 0.89<br>(0.12 to 1.83) | 899<br>(835 to 969) | 1058<br>(854 to 1280) | 153<br>(21 to 313) |
| Al-Mansoura | 10.57<br>(9.87 to 11.23) | 12.44<br>(10.04 to 15.04) | 1.65<br>(0.29 to 3.58) | 1807<br>(1688 to 1921) | 2127<br>(1717 to 2571) | 282<br>(49 to 613) |
| Abu Harbeh Aljadidah | 6.87<br>(6.38 to 7.38) | 8.09<br>(6.59 to 9.83) | 1.19<br>(0.10 to 2.29) | 1175<br>(1091 to 1262) | 1383<br>(1127 to 1681) | 204<br>(17 to 391) |
| Kod Al-Othmani | 0.31<br>(0.23 to 0.40) | 0.36<br>(0.26 to 0.49) | 0.05<br>(0.03 to 0.09) | 53<br>(39 to 69) | 62<br>(44 to 83) | 9<br>(5 to 15) |
| Al-Othmani | 1.76<br>(1.55 to 1.98) | 2.07<br>(1.63 to 2.53) | 0.31<br>(0.09 to 0.54) | 301<br>(265 to 339) | 354<br>(279 to 433) | 53<br>(16 to 93) |
| Al-Radwan | 27.31<br>(25.72 to 28.95) | 32.15<br>(26.41 to 38.71) | 4.98<br>(1.01 to 9.66) | 4670<br>(4398 to 4951) | 5498<br>(4516 to 6619) | 852<br>(172 to 1652) |
| Salah Al-Deen | 2.08<br>(1.86 to 2.30) | 2.45<br>(1.98 to 3.02) | 0.34<br>(0.06 to 0.68) | 356<br>(318 to 393) | 419<br>(338 to 517) | 58<br>(10 to 116) |
| Faqam | 0.81<br>(0.67 to 0.96) | 0.95<br>(0.73 to 1.18) | 0.14<br>(0.06 to 0.26) | 138<br>(114 to 164) | 163<br>(125 to 202) | 24<br>(10 to 45) |
| Al-Qateea' | 9.53<br>(8.9 to 10.16) | 11.22<br>(9.26 to 13.47) | 1.73<br>(0.24 to 3.34) | 1630<br>(1522 to 1738) | 1919<br>(1583 to 2304) | 295<br>(41 to 571) |

**Figure S13.**
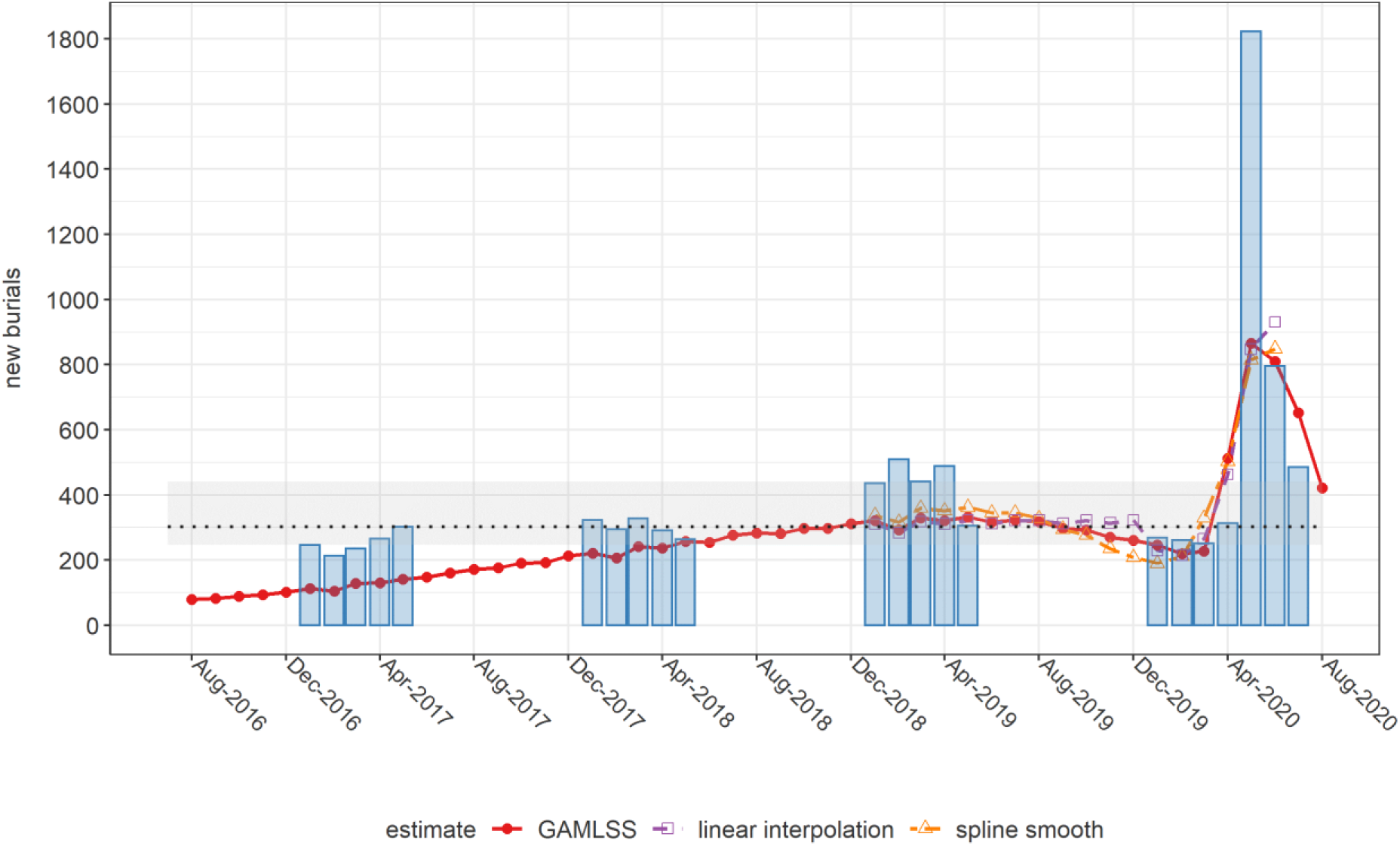
Comparison of monthly estimates of new burials across Aden governorate, by analysis method, and corresponding records from the Civil Registry office (blue columns). The horizontal dotted line and shaded area indicate the median, minimum and maximum monthly burials based on the 2017-2019 Civil Registry time series.

**Table S8.**
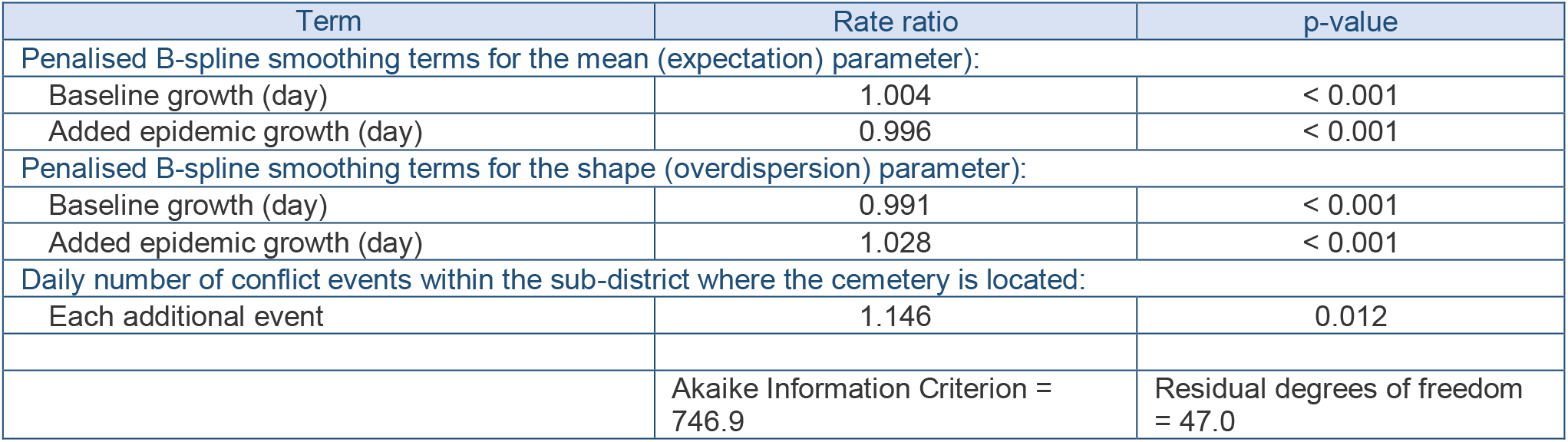
Fit statistics for an alternative GAMLSS model featuring the rate of insecurity events as an additional predictor. Coefficients are exponentiated to provide linear scale rate ratios.

| Term | Rate ratio | p-value |
| --- | --- | --- |
| Penalised B-spline smoothing terms for the mean (expectation) parameter): |  |  |
| Baseline growth (day) | 1.004 | < 0.001 |
| Added epidemic growth (day) | 0.996 | < 0.001 |
| Penalised B-spline smoothing terms for the shape (overdispersion) parameter): |  |  |
| Baseline growth (day) | 0.991 | < 0.001 |
| Added epidemic growth (day) | 1.028 | < 0.001 |
| Daily number of conflict events within the sub-district where the cemetery is located: |  |  |
| Each additional event | 1.146 | 0.012 |
|  | Akaike Information Criterion = 746.9 | Residual degrees of freedom = 47.0 |

**Figure S14.**
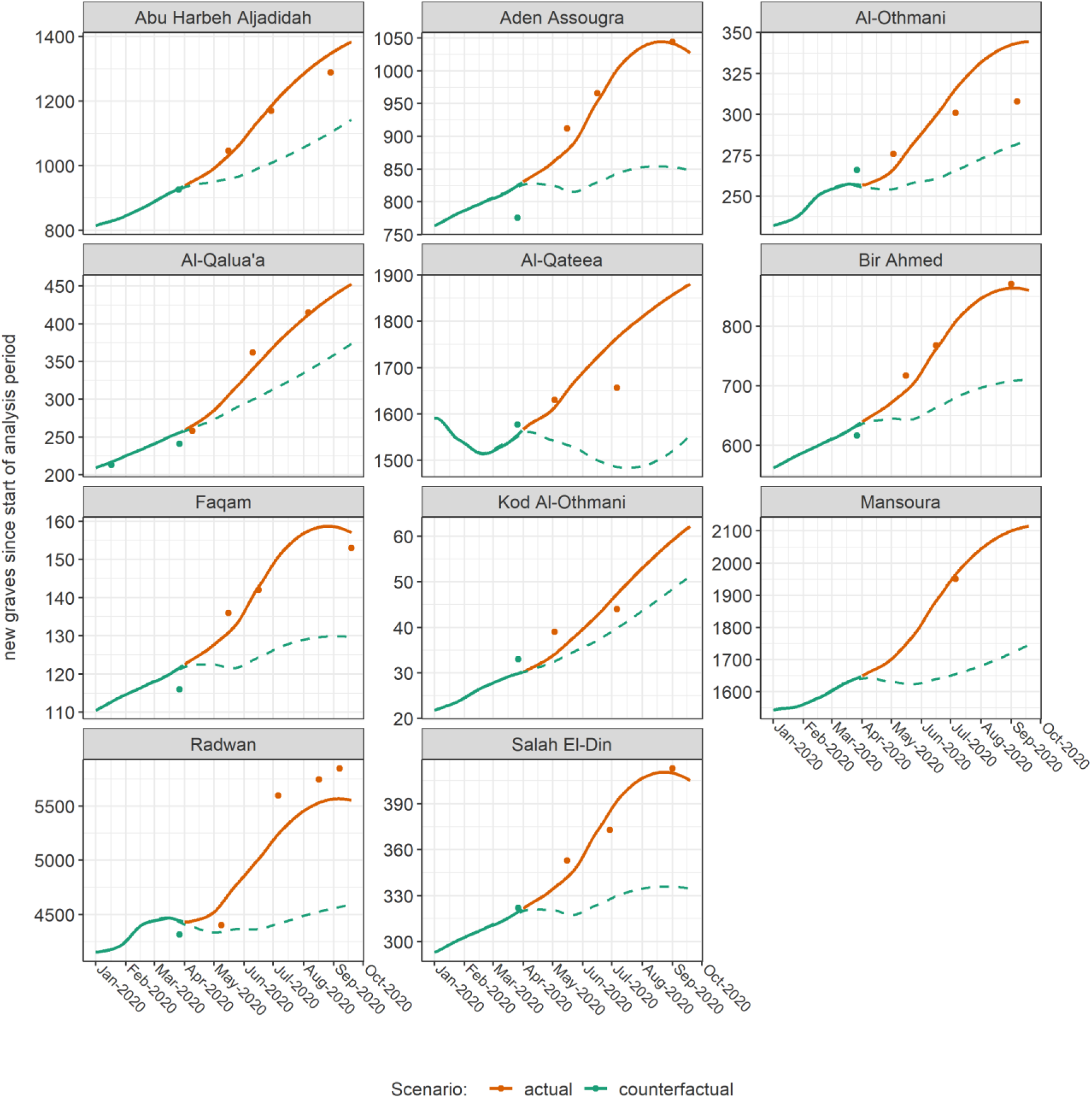
Predictions (lines) versus observations (dots), by cemetery, for the first 9 months of 2020, from an alternative GAMLSS model featuring the rate of insecurity events as an additional predictor. The dashed lines denote counterfactual predictions. Prediction lines have been moderately smoothed to avoid the jaggedness resulting from daily insecurity event fluctuations.

