## Supplementary material for "Excess mortality during the COVID-19 pandemic in Aden governorate, Yemen: a geospatial and statistical analysis": Frequently Asked Questions

26 October 2020

#### Who are you and who funded you to do this?

The study team comprises academics at the London School of Hygiene and Tropical Medicine (LSHTM), a post-graduate school of public health, and geospatial analysts at the Satellite Applications Catapult, a not-for-profit organisation. Both institutions are in the United Kingdom. Public health scientists at the University of North Carolina and Hadhramout University contributed expertise and data on a pro bono basis.

The study was funded by the United Kingdom Foreign, Commonwealth and Development Office (FCDO), through separate grants to the LSHTM and to the Satellite Applications Catapult. However, FCDO does not have any say in study design, analysis or interpretation of the findings, and has not interfered with the dissemination process. As such, this is an independent, scientific study whose principal aim is to help decision-makers in Yemen acquire better situational awareness of the pandemic and its impact. We have no political or advocacy agenda, and no conflicts of interest to declare.

#### You estimate that about 2000 people died of the virus in Aden, right?

No, not quite. We estimate that about 2000-2400 *excess burials* occurred in Aden governorate between April and September 2020. The word 'excess' means deaths above and beyond the 'baseline' level that might have been expected, based on patterns in previous months and years. However, we cannot say whether all these deaths are in fact due to COVID-19 disease. In fact, it's likely that some are due to the indirect effects of the pandemic, for example disruptions to health services (widely reported in Aden) or movement restrictions that would have kept other critically ill people from accessing care. In April 2020, major flooding affected Aden, and an increase in the incidence of malaria, dengue and chikungunya virus are also reported over our study period. It is thus possible that some of the excess is due to these pathogens (note however that floods and epidemics are recurrent in this region of Yemen, e.g. Aden was heavily affected by cholera in 2016-2018).

#### You counted individual graves. But how would you have picked up instances of people being buried together or cremated?

Ahead of data collection, we consulted a network of Yemeni researchers and civil society actors to understand local burial patterns, and whether these had been affected by the armed conflict or COVID-19. We don't believe multiple burials within the same grave, or mass burials, occurred in this region of Yemen during the study period. Cremation is inconsistent with religious beliefs and not practiced in the study setting.

### How does the level of mortality you found in Aden compare to that in high-income countries?

It’s very difficult to draw a straightforward comparison. Superficially, the Aden results indicate that at the peak of the epidemic, excess deaths were on a similar scale to those seen in New York, London, Paris and Madrid. However, in European and North American cities, the first wave of the pandemic peaked much earlier than it would have in the absence of the very stringent lockdowns that Europe and North America implemented: thus, the excess mortality figures from the first wave in high-income countries don’t reflect what might have happened in the absence of control. In Aden, it doesn’t appear from information available to us that control measures were of sufficient stringency to suppress virus transmission (i.e. drive the R index below 1). We thus believe it’s more likely that what we are observing in Aden is the ‘natural’ peak of the epidemic, or at least of its first wave.

### So you’re saying that the COVID-19 pandemic has been less severe in Aden than in high-income countries?

Not necessarily. First of all, our study has a number of limitations that we consider in the paper. Secondly, we need to consider another key factor, namely age: Aden has a much younger population than, say, Madrid, and this difference in age distribution would, all else being equal, result in lower mortality per capita in Aden. In fact, mathematical models developed by the LSHTM back in June 2020 predicted a per capita level of mortality due to COVID-19 throughout Yemen that is broadly similar to the one we estimate for Aden. Those mathematical models took into account Yemen’s age distribution and also what is known about the extent to which different age groups mix with each other, an important parameter that varies from setting to setting. However, in other respects the models were based on what was known about the virus from China and Europe. In short – if we adjust for known differences in key parameters between Aden and, say, Madrid, our estimate is broadly in line with what we would have expected in a scenario with limited effective control.

### Can these results be extrapolated to the rest of Yemen?

With caution. It’s fairly plausible that in other urban settings in Yemen, where control measures were similar or less stringent, the epidemic would have unfolded with a broadly comparable pattern. However, in rural parts of the country we assume that transmission would be lower (due to less overcrowding), leading to slower, more protracted epidemic curves.

### What do you results mean in terms of the progression of the pandemic in Yemen?

It’s difficult to make a sweeping statement about Yemen, but for Aden at least, and perhaps by extension for other urban settings in Yemen, we think these findings indicate that the first wave of the COVID-19 epidemic has mainly passed. We think that this did not occur because control measures suppressed transmission, but because enough people were infected that the population reached herd immunity, a point in epidemic when so many people are at least temporarily immune that transmission starts to decline on its own. Of course, control measures and behaviour change may have also helped. Obviously, our study is not the optimal source of information on the evolution of the epidemic. A well-conducted seroprevalence survey (i.e. a study that estimates the proportion of people in the population who carry antibodies suggesting prior infection) would provide a more conclusive picture of the situation.

#### So you're saying that the epidemic is over in Yemen? Should Yemen then consider itself rid of the virus, and return to normal life?

No, we can't say that. We think it's plausible that in Aden at least, the population might have acquired temporary herd immunity, meaning they would be protected for a while from a second pandemic wave. Anecdotal reports and surveillance data from elsewhere in Yemen broadly also suggest that the epidemic has waned. However, we really don't know at this point how long immunity lasts. It could be short-lived, meaning that the Yemeni population could be vulnerable to new waves of the pandemic in the not-so-distant future. Moreover, it isn't as if the virus simply goes away when herd immunity is reached. Usually, it continues to circulate at very low levels in pockets of the population, or gets reintroduced from abroad. Ongoing surveillance is thus imperative.

#### The satellite imagery method you used seems to undercount burials, particularly the farther you go back in time. Doesn't this undercount bias your baseline mortality estimate?

Yes, there seems to be considerable bias with our method when applied to images from 2-3 years ago – at least if we compare the number of burials it yields with data from civil registration offices in Aden, or plausible overall death rates. We are working on how to improve our geospatial analysis techniques to deal with this problem. In the event, the statistical model we used to develop a baseline death rate is inherently much more guided by recent data points, so we are confident that the above bias won't have affected our findings appreciably.

#### Still – do you really think this satellite imagery method is a viable option?

Yes. It's one of the only options we can think of for places where ground access is restricted. It's potentially quite efficient, and can be replicated again and again over time, to provide some sort of real-time monitoring. Moreover, if applied prospectively, the method would rely on newer-generation satellites and would have the option of commissioning bespoke imagery, which would come with the right specifications and resolution – thereby probably improving our ability to detect individual graves.

#### Yes, but is it really ethical for you to analyse satellite images of people's lives without them being aware or consenting?

The study was approved by the ethics committee of the LSHTM. The satellite images we used are commercially available – they are collected by satellites on a regular basis, and then sold to anyone who wishes to purchase them (see the paper). Satellite images come with different resolution. We did not have access to the kind of resolution that militaries or intelligence agencies might use: in the imagery we used it's completely impossible to distinguish human beings. We are, however, aware that even satellite imagery analysis carries issues of research ethics and integrity. We don't think these are yet very well worked out for the application of estimating mortality, mainly because there is really little precedent of this use, as far as we are aware. Therefore, we are planning to reflect on this case study, with the help of ethics experts, and more fully consider the ethical dimensions of this kind of work, so as to inform future application of this method.
