## Supplementary material for "Excess mortality during the COVID-19 pandemic in Aden governorate, Yemen: a geospatial and statistical analysis": Abstract in Arabic

### زيادة في عدد الوفيّات خلال جائحة كوفيد -19 في محافظة عدن، اليمن:
### تحليل جيومكاني وإحصائي


**ملخّص البحث:**

**المقدّمة:**

إنّ عبء جائحة كوفيد-19 في البلدان منخفضة الدّخل وتلك المتضرّرة من النّزاع لا يزال غير واضح ، وهو يعكس إلى حدّ كبير نسباً منخفضة لإجراء اختبارات الكشف عن الفيروس . بلغت أعداد حالات دخول المستشفيات وعمليّات الدّفن ذروتها خلال الفترة بين أيّار (مايو) وحزيران (يونيو) من عام 2020، وذلك وفقاً للتّقارير الصّادرة من بعض أنحاء اليمن . وبهدف تقدير الزّيادة في عدد الوفيّات التي حدثت خلال فترة الوباء ، قمنا بقياس نشاط أعمال الدّفن بشكلٍ كمّي في جميع المقابر التّي أمكن تحديدها داخل محافظة عدن في اليمن (حيث أنّ عدد السّكّان يقدّر بحوالي مليون نسمة) ، وذلك من خلال تحليل صور الأقمار الصّناعيّة ذات الدقّة العالية جدّاً ومن ثمّ مقارنة التّقديرات النّاتجة مع بيانات مكتب السّجلّ المدنيّ في المدينة .

**طرق البحث:**

قمنا بجمع معلومات ميدانيّة وأخرى عن بعد لتحديد المقابر التّي شهدت نشاطاً ملحوظاً في أعمال الدّفن في محافظة عدن، ومن ثمّ قمنا بتطبيق تقنيّات التّحليل الجيومكاني بهدف تحديد القبورالحديثة بطريقة يدويّة أولاً وقياس التّغييرات في مساحة السّطح المستخدَم للدّفن ثانياً على مدار فترةٍ زمنيّة بدءاً من تمّوز (يوليو) عام 2016 إلى أيلول (سبتمبر) عام 2020 . تمّ احتساب عدد القبور الفائتة (أي التّي لم تلتقطها صور الأقمار الصّناعيّة) من خلال تحليل بيانات مساحة السّطح المستخدم للدّفن . ومن ثمّ استخدمنا أساليباً بديلة تتضمّن الاستنباط (الاستقراء) البسيط ونموذج النّمو المختلط المضاف المعمّم ، للتّنبّؤ بمعدّلات الدّفن الفعليّة والافتراضيّة (أي في حال عدم وجود الوباء) ، لكلّ مقبرةٍ وللمقابر في كامل المحافظة وذلك خلال الفترة الأكثر احتمالًا لحدوث وفيّات بسبب كوفيد-19 (أي بدءاً من الأوّل من نيسان (أبريل) عام 2020) . تمّ فيما بعد حساب الزّيادة في عدد عمليّات الدّفن . قمنا أيضًا بتحليل إشعارات الوفاة المقدّمة إلى مكتب السّجلّ المدني خلال الفترة بين نيسان و تمّوز(أبريل ويوليو) من عام 2020 وفي السّنوات السّابقة .

**النّتائج:**

قمنا بجمع 78 مشاهدة (رصد) من 11 مقبرة وتوجّب استخدام طريقة التّغير في مساحة السّطح المستخدم للدّفن لاحتساب عدد القبور في عشر مقابر منها . في البدء تراوحت أحجام المقابر من حيث عدد القبور بين 0 إلى 6866 قبراً . كان هناك ارتفاعٌ واضحٌ في معدّلات الدّفن اليوميّة في عشر مقابرمنها ووصل إلى ذروته في الفترة بين نيسان (أبريل) و تموّز (يوليو) من عام 2020 في عشر مقابر . قدّرت طريقتا الاستنباط (الاستقراء) والنموذج المختلط المستخدمتين زيادةً بمقدار ≈ 1500 مدفن حتى تاريخ 6 تموّز(يوليو) و 2120 مدفنٍ حتى تاريخ 19 أيلول (سبتمبر) ، وهو ما يعادل ذروةً في الوفيّات الأسبوعيّة تبلغ نسبة 230٪ من المعدّل الافتراضي (أي في حال عدم وجود الوباء) . كانت تقديرات صور الأقمار الصّناعيّة للزّيادة في الوفيّات أقلّ من بيانات مكتب السّجلّ المدني بشكلٍ عام ، وقد أشارت الأخيرة إلى ذروةٍ في عدد الوفيّات وصلت إلى 1823 حالة وفاة في شهر أيّار (مايو) منفرداً. ومع ذلك ، فقد أشار كلا المصدرين إلى أنّ الوباء قد تراجع بحلول أيلول (سبتمبر) من عام 2020.

**المناقشة:**

تعتبرهذه الدّراسة الأولى من نوعها ،على حسب علمنا، في استخدام صور الأقمار الصناعيّة لتقدير معدّل وفيّات السّكّان. تشير النّتائج إلى أن جائحة كوفيد-19 في هذه المحافظة اليمنيّة الحضريّة تحمل أثراً كبيراً لم يتمّ الإلمام بكافّة جوانبه بعد. كما أنّ النّتائج تتطابق عموماً مع تنبّؤات النّماذج الرّياضيّة السّابقة لمسار الجائحة في اليمن . من الجدير بالذّكر أنّ الطريقة المتّبعة هنا غير قادرةٍ على التّفريق بين الوفيّات النّاجمة عن فيروس كوفيد-19 بشكلٍ مباشر وتلك غير المباشرة . يمثّل تحليل صور الأقمار الصّناعيّة للمقابر نهجًا مبتكراً وواعدًا لرصد الأوبئة وآثار الأزمات الأخرى، لا سيّما عندما يكون من الصّعب جمع البيانات بشكلٍ ميدانيّ .
